# Protocol for an interdisciplinary cross-sectional study investigating the social, biological and community-level drivers of antimicrobial resistance (AMR): Holistic Approach to Unravelling Antibiotic Resistance in East Africa (HATUA)

**DOI:** 10.1101/2020.12.19.20248543

**Authors:** Benon B. Asiimwe, John Kiiru, Stephen E. Mshana, Stella Neema, Katherine Keenan, Mike Kesby, Joseph R. Mwanga, Derek J. Sloan, Blandina T. Mmbaga, V Anne Smith, Stephen Gillespie, Andy G. Lynch, Alison Sandeman, John Stelling, Alison Elliott, David M. Aanensen, Gibson S. Kibiki, Wilber Sabiiti, Matthew T. G. Holden, HATUA CONSORTIUM, Catherine Kansiime, Martha F. Mushi, Arun Gonzales Decano, Dominique L. Green, John Mwaniki, Nyanda E Ntinginya, Joel Bazira

## Abstract

**Introduction:** Antimicrobial resistance (AMR) is a global health threat that requires urgent research using a multidisciplinary approach. The biological drivers of AMR are well understood, but factors related to treatment-seeking and the social contexts of antibiotic (AB) use behaviours are less understood. Here we describe the Holistic Approach to Unravelling Antibacterial Resistance in East Africa (HATUA), a multi-centre consortium that investigates the diverse drivers of drug-resistance in urinary tract infections (UTIs) in East Africa.

**Methods and Analysis:** This study will take place in Uganda, Kenya and Tanzania. We will conduct geospatial mapping of AB sellers, and conduct mystery client studies and in-depth interviews (IDI) with drug sellers to investigate AB provision practices. In parallel, we will conduct IDIs with doctors, alongside community focus groups. Clinically diagnosed UTI patients will be recruited from healthcare centres, provide urine samples, and complete a questionnaire capturing retrospective treatment pathways, socio-demographic characteristics, attitudes and knowledge. Bacterial isolates from urine and stool samples will be subject to culture and antibiotic susceptibility testing (C&AST). Genomic DNA from bacterial isolates will be extracted with a subset being sequenced. A follow-up household interview will be conducted with 1800 UTI-positive patients, where further environmental samples will be collected. A sub-sample of patients will be interviewed using qualitative tools. Questionnaire data, microbiological analysis and qualitative data will be linked at the individual level. Quantitative data will be analysed using statistical modelling including Bayesian network analysis, and all forms of qualitative data analysed through iterative thematic content analysis.

**Ethics and Dissemination:** Approvals have been obtained from all national and local ethical review bodies in East Africa and the UK. Results will be disseminated in communities, with local and global policy stakeholders, and in academic circles. They will have great potential to inform policy, improve clinical practice and build regional pathogen surveillance capacity.

## Introduction

Antimicrobial resistance (AMR) emerges when pathogens evolve ways to survive treatments (i.e. antibiotic, antiprotozoal, antiviral and antifungal medicines). Antibiotic resistance (ABR) is a significant subset of this phenomena and is the focus of this study. Increasing levels of resistance to antibiotics^1^ are a serious threat to global health, and, if no action is taken, are projected to cause 10 million excess deaths by 2050 [1]. It is unlikely that this burden will be evenly distributed and Africa is particularly vulnerable to the challenges posed by AMR since the continent suffers the highest morbidity and mortality arising from infectious diseases and the least developed laboratory infrastructure [2]. The economic, cultural and ethnic diversity of Africa mean that the problems surrounding drug resistance are likely both to be distinct from other regions of the world, and to display significant intra-continental diversity. Regional solutions and local approaches are necessary.

Beyond the microbiological and biological origins of ABR and AMR there are bio-social problems requiring an interdisciplinary approach that incorporate social science perspectives on human-microbial interactions [3]. Despite this, social science perspectives on the evolution and control of AMR are rare [4]. Whilst the biological drivers of AMR in pathogens are well explored[5], the extent to which these are modulated by human behaviour in and around antibiotics (AB) is less well understood. Cultural, social, economic and clinical factors play a part in shaping the way people source, consume, use and distribute antibiotics (AB) [6]. More effective AB stewardship is acknowledged as one of the key interventions to preserve existing ABs. If this is to be achieved, then we must address the knowledge gap of structural factors and social behaviours that drive pathogens’ antibiotic selection pressure and ultimately genetic changes in pathogens. Such a problem cannot be achieved by one scientific discipline acting alone. Rather, this complex, multifaceted problem requires an integrative approach able to work effectively across disciplines.

Here, we describe a newly formed consortium – the Holistic Approach to Unravelling Antibacterial Resistance (HATUA). The consortium brings together expertise in microbiology, pathogen genomics, epidemiology, human geography, anthropology, sociology, computational biology and statistics across 7 institutions, from three East African countries (EAC), the UK and the United States. Taking ‘*Hatua*’ (a Kiswahili word for ‘step’ or ‘action’) as inspiration for its acronym, the consortium addresses the social and biological drivers of antibiotic drug-resistance in multiple sites in Kenya, Tanzania and Uganda, using the clinical prism of urinary tract infection (UTI). UTIs are common globally, and, in LMICs rarely have a laboratory diagnosis. Moreover, they are often mistaken for other illnesses such as sexually transmitted infections (STIs), and consequently remain poorly treated [7,8]. Through synthesis across multiple sites our study, of the burden and drivers of AMR at national and regional levels, will provide insights on ABR emergence that may be applicable to other diseases and contexts.

The research will focus on four key elements of the ABR problem in UTI: the therapy landscape, the pathogen, the patient, and the community. The research will target three main (inter-related) drivers: First, the supply of antibiotics, second, the level of knowledge of proper use of antibiotics and third, the choice of AB by clinicians and patients and relative effectiveness of treatment. In each case, we will bring novel methodological and theoretical approaches to bear on the issues. Our work will deliver a unique research dataset that links patients, the pathogen, and the socio-economic and socio–demographic picture of the individual and their community. Through our research and impact activities, we will also strengthen diagnostic and analytical capacity, and dynamic pathogen surveillance in the region.

### Theoretical and conceptual framework

A challenge of inter-disciplinary working is not necessarily finding commonly understood methodologies but shared theoretical (ontological and epistemological) frameworks. This is a theme with which the ‘one health’ paradigm must grapple if it wishes to understand how infectious disease processes are products of both biological and social relations [9]. In this study, we conceptualise the drivers of AB-resistant UTI infections as part of a complex system of interrelating biological and social entities [10], drawing theoretical inspiration from assemblage theory, which facilitates the incorporation of a range of different material actors/actants (humans, animals, microbes) in a single dynamic system [11]. Its advantage is that it encourages experimentalists to engage with the conscious human decision-making in bio-social systems. This approach encourages social scientists to take an ‘ontological turn’, recognising that ‘inanimate’ material things (be they bacteria, drugs, clinics etc), are not merely inert unless given meaning by human subjects, but are themselves able to animate and produce effects. [12] Consequently, we argue that ‘new materialist’ approaches[13] provide an ideal framework for conceptualising AMR as a complex assemblage of human and non-human entities operating at various scales.

In developing our study we reviewed the extant literature on possible social and biological drivers and constructed a representation of a UTI-related AMR assemblage (see simplified version Fig.1). The representation maintains the false poles of ‘social’ and ‘biological’ but only to emphasise the importance of exploring the integration of socio-biological factors that facilitate AMR. We know that AB suppress bacteria exerting selection pressure that results in the development of ABR (Fig. 1, blue/right) but some of the drivers of this process lie in the social realm (Fig 1, white/left). The behaviours of individual agents such as health care workers, patients and those in the community are situated within the structures and institutions that shape them. For example, improvements in structural inequalities around water, sanitation and hygiene (WASH) at the household or community-scale might reduce the development of UTIs and the need for antimicrobial use in the first place [14], removing one significant driver of AMR selective pressure.

**Figure 1:**
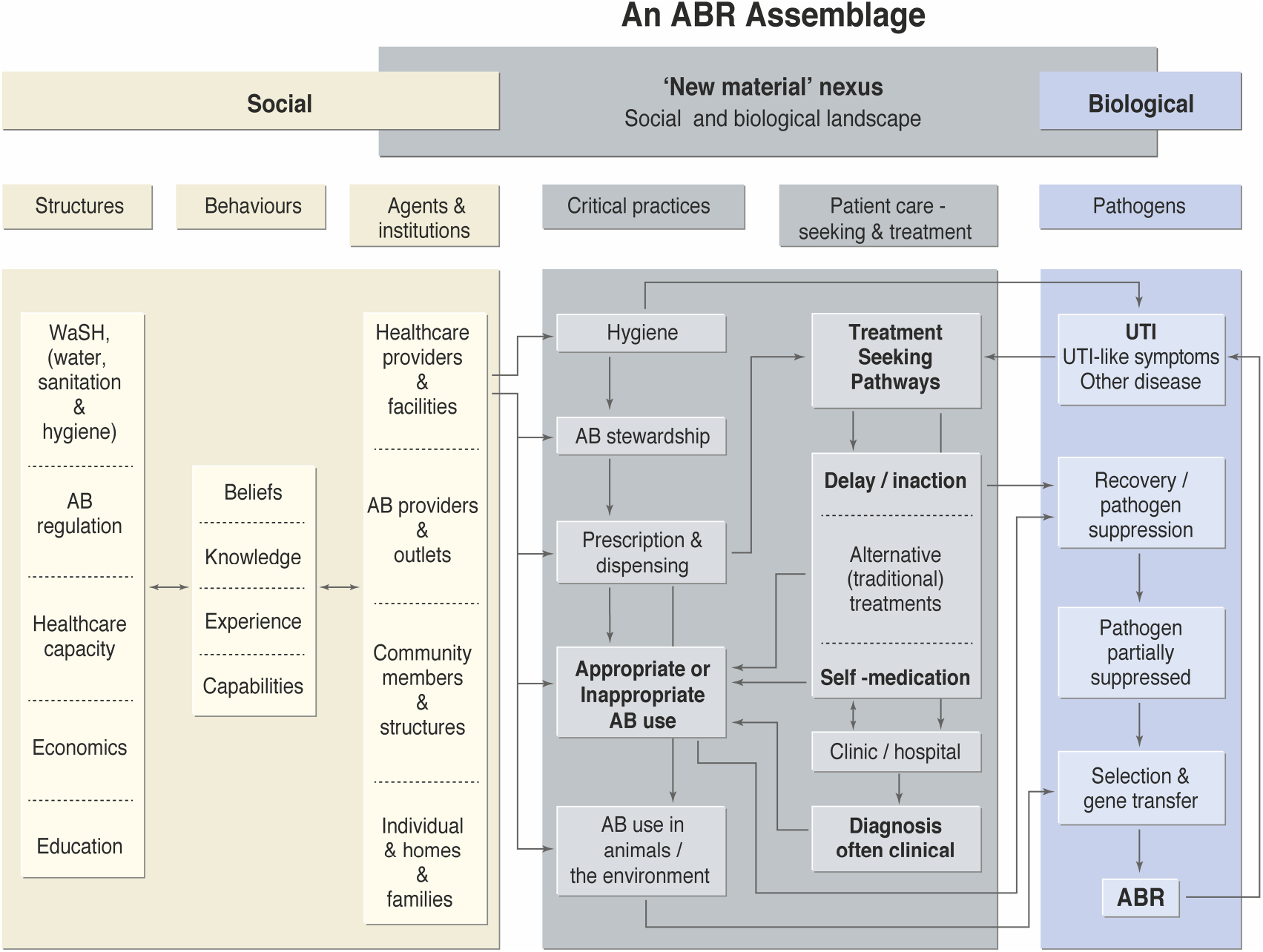
An ABR assemblage (a complex set of inter-related factors) as it refers to UTI.

Previous research has focused on challenges faced by clinicians in attempting to prescribe antimicrobials effectively, such as knowledge, diagnosis, and drug availability [15]. However in EAC, pharmacies, drug shops and other informal sellers are ubiquitous, existing side by side with the public and private healthcare systems, other providers such as traditional healers, and veterinary providers. With such medical pluralism, non-prescription dispensing of antimicrobials for self-medication is extremely common [16,17], influenced not only by individual-level abilities but by larger social and regulatory structures.

At the centre of this assemblage we identify a ‘new material’ nexus (coloured grey) consisting of critical practices (such as dispensing, AB use in animals and AB stewardship practices), and the treatment-seeking pathways of UTI patients. This central nexus describes the entanglement of socio-biological processes that likely drive selective pressure in bacteria and thence development of AMR UTI. A focus for HATUA is to describe and investigate the social and biological drivers of patients’ treatment seeking pathways, or “patient pathways”, and how these relate to patterns of AMR at individual and community level. We conceptualise a patient pathway as a longitudinal sequence of health seeking behaviours taken by individuals when they feel ill, which might include delays in seeking treatment, self-medication, [18], attending various formal and informal healthcare providers, and taking medications more or less appropriately. These pathways may be complex, non-linear and, given the economic and socio-cultural barriers to clinically ideal pathways, iterative. Rather than theorise healthcare decision making as patients freely and rationally choosing from a suite of available options, pathway-based models view behaviour as a sequence of steps each with its own situated rationality, and set of social dependencies, constraints and inter-relationships [19,20]. This draws on concepts of ‘medical syncretism’ which describe how patients may oscillate between different types of healthcare in a single illness episode. [21] Requiring detailed longitudinal analysis, this approach has most often been operationalised using qualitative interviews, as in the case of abortion-related care [22–24] and quantitative approaches are rare [25–27]. In this study we collect large-scale quantitative data alongside qualitative IDIs, which are linked to individual-level pathogen and AMR profiles.

### Pilot Phase 2017-18

HATUA pilot activities in 2017-18 in Uganda demonstrated the feasibility of the holistic approach by conducting a study of microbiological and genomic features of urinary pathogens collected from clinic patients, combined with quantitative socio-demographic data. Genomic characterisation of the strains (predominantly *E. coli* and *K. pneumoniae*) revealed high levels of resistance mainly disseminated via clonal and horizontal transfer [28]. We also conducted in-depth interviews with patients and healthcare providers, and focus group discussions with community members in Mbarara district, Uganda to explore behaviours and attitudes to AB use. These highlighted potential drivers of AMR including distrust and misuse of ABs, failure to complete treatment courses, human use of veterinary drugs and combined consumption of ABs and traditional medicines, which informed the development of the main study.

## Methods and Analysis: Main HATUA study

### Field Activities and Data Collection

HATUA’s activity will take place in Kenya, Uganda and Tanzania and multidisciplinary teams will sequentially survey in three study areas (SA) in each country (Figure 2). The locations are socio-demographically distinct: (1) Urban, economically advanced settings that potentially increase affordability and access to ABs; (2) Remote villages in poorer areas, where poverty, physical isolation and lower access to ABs possibly lead to potential drivers such as sharing of prescriptions, restricted microbiological culture and poor AB susceptibility testing (C&AST) capacity; and (3) Pastoralist and neglected network - areas with highly mobile pastoralist communities, and high levels of animal-human interaction fostering a zoonotic link. The tertiary hospital (level 5), and lower level healthcare facilities in the SAs (Level 4, 3 and 2 clinics/ hospitals) will be used to recruit UTI patients and also conduct interviews with healthcare providers (see Work streams (WS) 1, 2 and 3 below). Beyond the hospitals, the activity will take place in the rest of the SA, visiting AB retailers, households and communities. At the start of activities in all SAs, community inception workshops will be held to introduce and communicate the goals of HATUA to relevant stakeholders (the local community, village health teams, doctors, hospital and lab workers, local health authorities). Sampling will begin in April 2019 in some sites and will continue through 2020 (Covid-19 permitting). All data collection tools and operating protocols are standardised allowing valid comparison across sites and countries. For collecting quantitative, social science and laboratory data, and geospatial mapping we will use EpiCollect5 (https://five.epicollect.net) [29] a customisable mobile data gathering tool installed on tablets and mobile phones.

**Figure 2:**
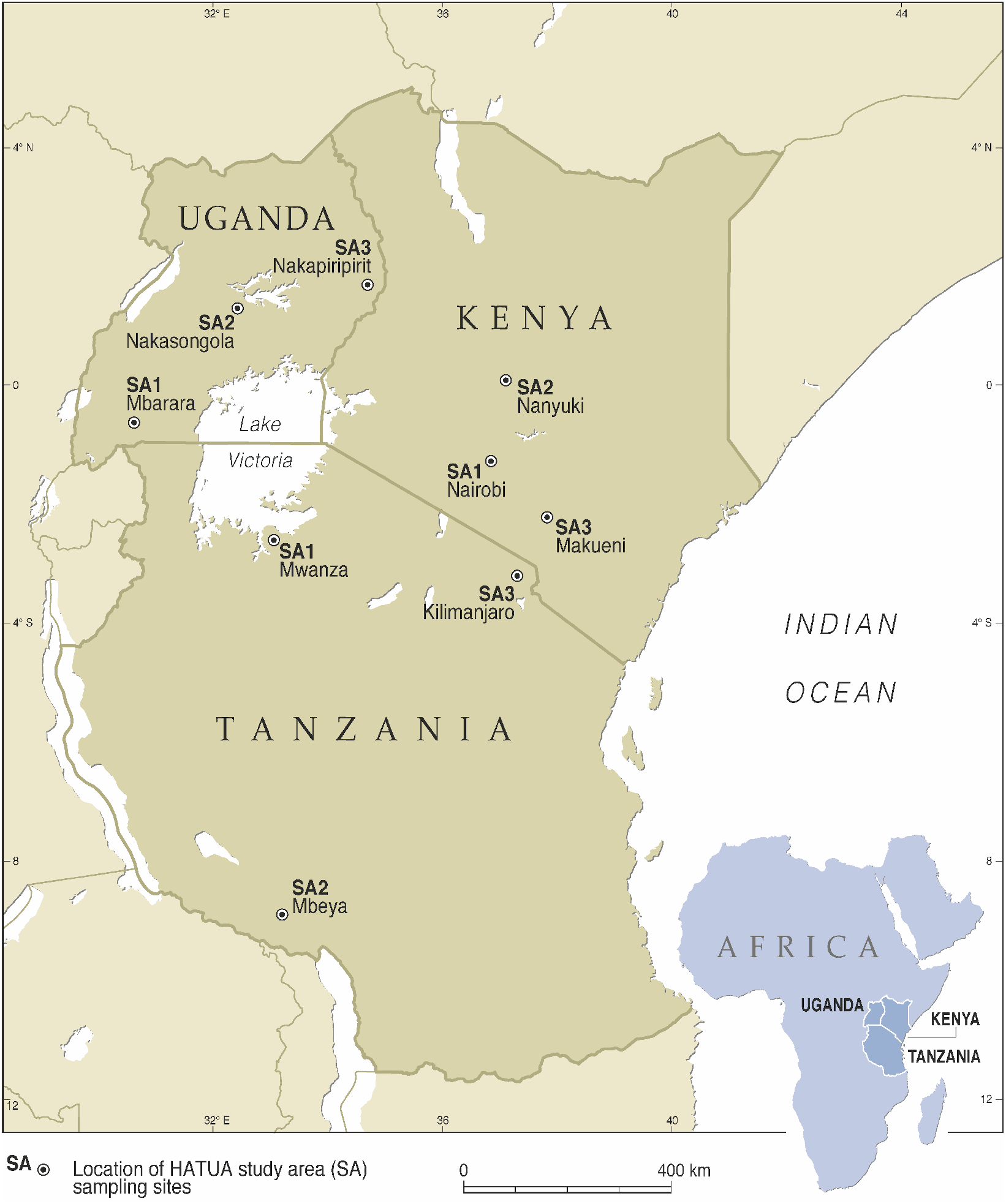
Location of HATUA Study Areas (SAs)

### Recruitment of sample of UTI Patients

At the heart of the HATUA study will be a linked data set of 1800 patients (600 per country, 200 per site with culture-confirmed UTI (Table 1). Given the estimated UTI prevalence from the pilot study, this will mean recruiting three times that number, c. 5400 patients. Sample size calculations were challenging, given the number of possible ways of measuring this, and limited evidence of different prevalence rates for community and hospital-acquired resistant UTI infections adults and children in this region. To estimate precision, under a binomial model, the numbers required to obtain a 95% confidence interval for the prevalence of 0.5 with width no greater than 0.1 would be a little under 400 (384). That model relies on there being no underlying population or sampling structure and so will lead to an underestimate of the true required numbers in our complex study. Our larger study size of 600 per country will provide some robustness to our ability to estimate this parameter with the desired accuracy, while allowing us to uncover some of the population structures that, if modelled correctly, will improve the precision in our estimate of prevalence. In level 2, 3, 4 and 5 hospitals in each SA we will recruit adult and child outpatients (min. 90% of the total sample), that a doctor identifies as suffering with UTI-like symptoms (e.g. burning /irritation during urination, dysuria, pyuria). In level 5 hospitals we will also recruit inpatients (max. 10% of the total). For non-pregnant child patients aged under 18 years data, will be provided by an accompanying parent or guardian. Our sample is representative only of the population of clinic attendees, rather than the general population and is likely to include a higher proportion of patients with treatment failures, who are wealthier, and patients living closer to clinics. However, clinic attendees are an important patient subset as these are the individuals specifically for whom clinicians must make patient management and treatment decisions.

**Table 1:** Target sample sizes for HATUA data collection tools.

|  | Per study site |  |  | Per country<br>TOTAL | Study<br>TOTAL |
| --- | --- | --- | --- | --- | --- |
|  | *Phase 1* | *Phase 2* | *Total* |  |  |
| **Patient questionnaire** | ~300, to recruit 100 UTI + | ~300, to recruit 100 UTI + | ~600, to recruit 200 UTI + | ~1800, to recruit 600 UTI + | ~5400, to recruit 1800 UTI+ |
| **Household questionnaire** | 100 | 100 | 200 | 600 | 1800 |
| **Patient IDIs** | 5 | 5 | 10 | 30 | 90 |
| **Healthcare worker IDIs** | 1-5 | 0-5 to fill quota | 5 | 15 | 45 |
| **Drug seller mystery client study visits** | - | 50 | 50 | 150 | 450 |
| **Drug seller IDIs** | - | 10 | 10 | 30 | 90 |
| **Focus group discussions** | - | 4-8 | 4-8 | 12-24 | 36-72 |

A urine sample and where possible a faecal sample will be taken from all patients. From catheterised inpatients urine catheter samples will be gathered, whereas outpatients will be advised how to self-collect mid-stream urine samples. In addition, patients will have a questionnaire administered to collect retrospective data on their recent clinical history, and related treatment seeking and AB usage (see Fig. 3). The questionnaire will also capture individual level socio-demographics (e.g. age, gender, education, household and family circumstances), household socioeconomic factors (e.g. housing type, amenities and asset ownership used to derive multidimensional poverty indices), attitudes, related behaviours and residential geographic information.

**Figure 3:**
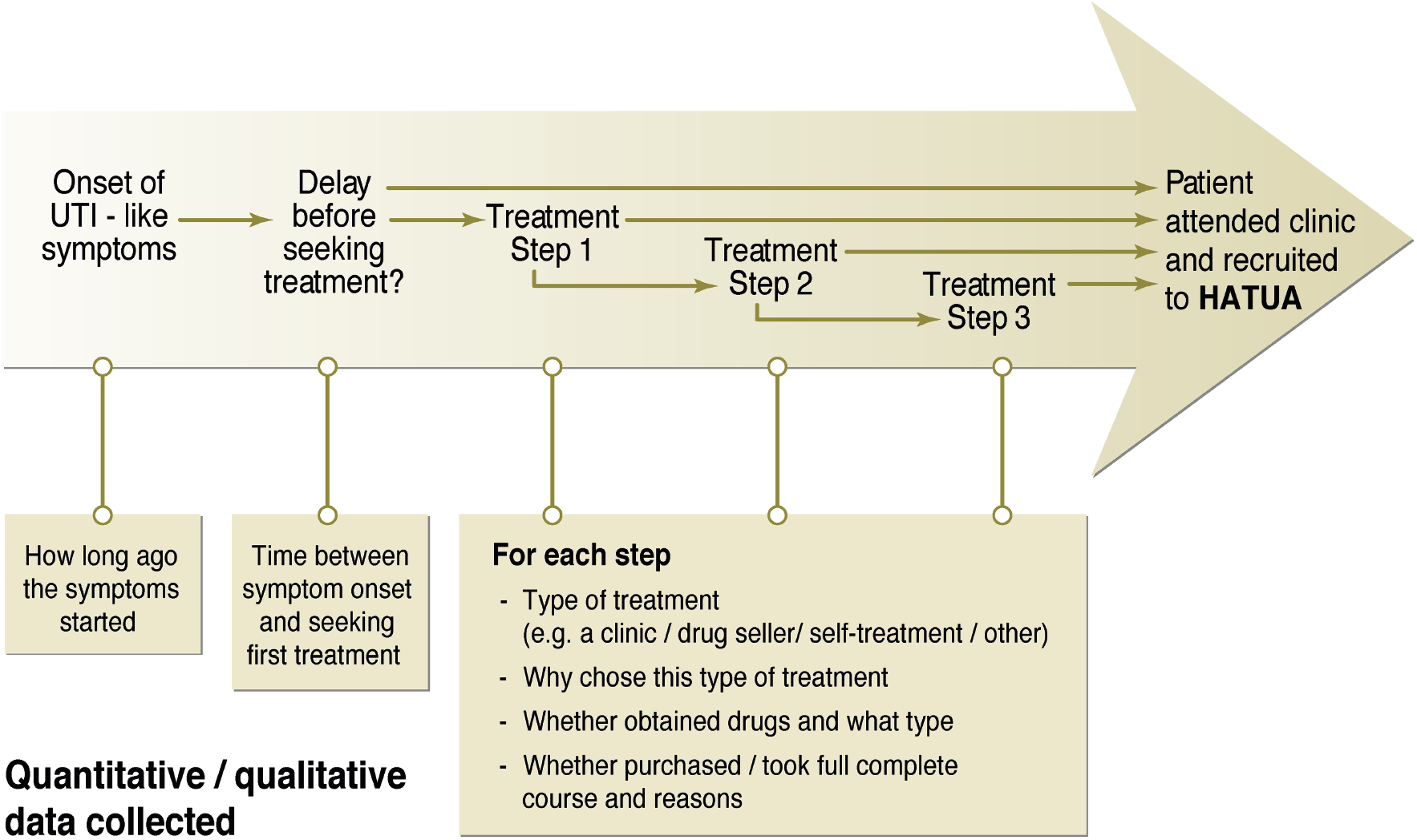
Description of quantitative and qualitative data collected about the self-reported’patient pathway’ in the linked patient sample.

During initial recruitment, all patients will be asked to consent to being followed up if they test positive for a UTI. Eligible outpatients with culture-confirmed UTI will be re-contacted for a follow-up to their homes (see WS4; below). For logistical reasons, only patients living within an approximate distance of 70 km from the level 5 hospital and 10 km from the level 4 and 3 hospitals will be eligible for follow up. During follow-up a questionnaire will be administered to a competent adult member of the household to capture sanitation and hygiene, socio-demographics, economic and poverty dimensions, household health seeking behaviour, and livestock keeping practices. Environmental sampling of soil, animal faecal samples and other materials in the immediate proximity of the home will be conducted. To enrich the quantitative data, a sub-sample of UTI patients will be purposively selected for qualitative IDIs based on their having drug-resistant UTI pathogens or reporting complex patient treatment pathways (10 per SA, 90 in total).

The resulting qualitative and quantitative social science data and microbiological data will form an individual-level linked dataset. This will incorporate quantitative questionnaire data collected at the clinic and the home, qualitative data from IDIs, C&AST and WGS of pathogens from the patient, and C&AST from household samples (see Fig. 4). This can be related to multi-scalar data on landscape (WS1).

**Figure 4:**
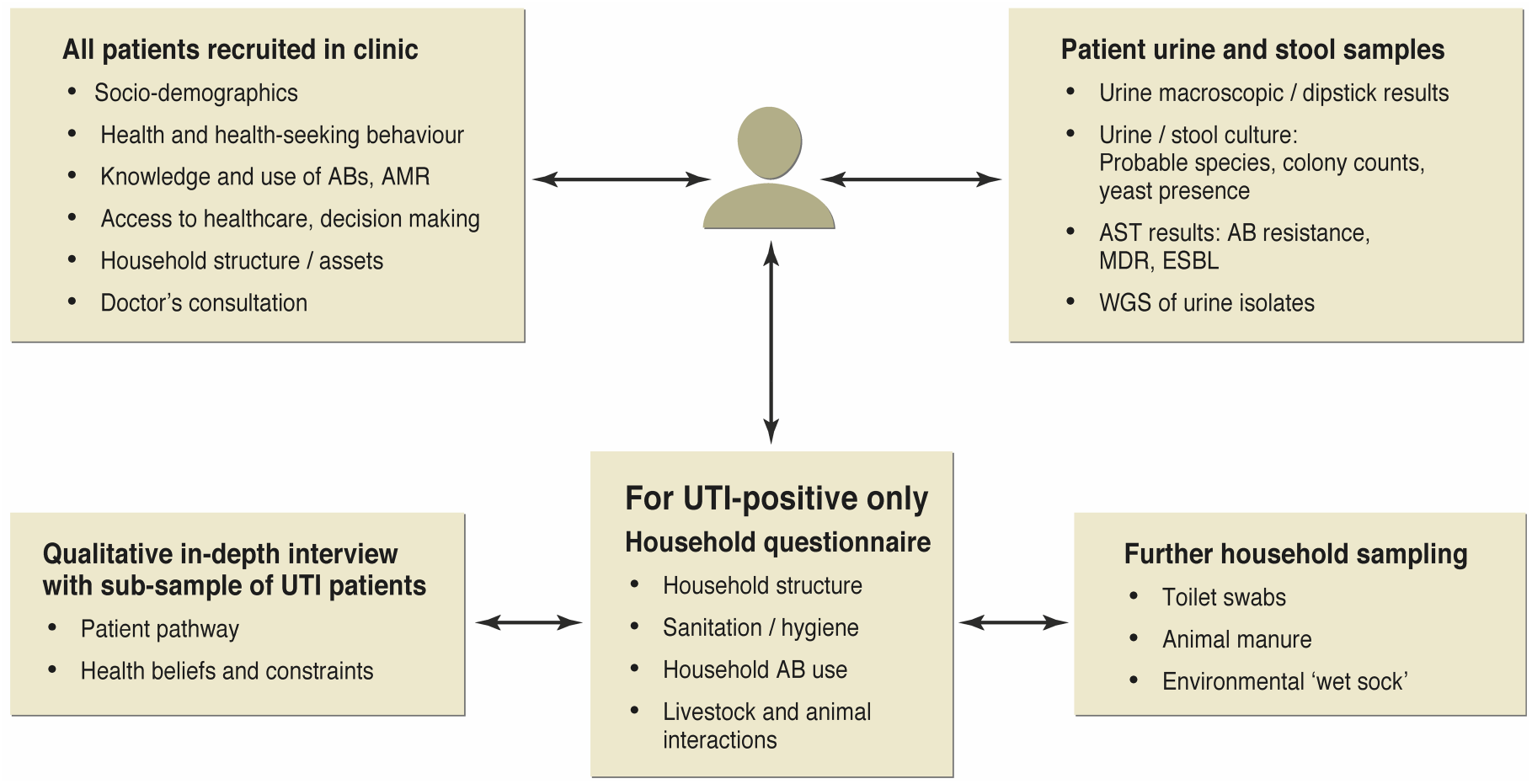
The linked individual-level patient dataset collected in HATUA.

### Further data collection

#### Geospatial mapping

Using EpiCollect on a GPS-enabled tablet, in all SAs, we will conduct geospatial mapping of observed AB providers in the local community (e.g. from hospitals and clinics, to retail pharmacies and informal drug sellers, to veterinary drug shops).

#### Mystery client study of drug sellers

A common means of reducing response bias from surveys is to use mystery client or simulated client studies to investigate ‘real life’ dispensing practices of drugs sellers and pharmacies [17,30]. Using the sampling frame created by the geospatial mapping exercise above, we will randomly select outlets to participate in a simulated/mystery client study. Trained fieldworkers using predefined scenarios will request ABs and/or advice for the treatment of UTI-like symptoms. After the encounter, they will record data including whether and what kind of AB they were offered, the course and regimen they were sold, whether they were asked for a prescription, costs, and advice given.

#### Qualitative IDIs with drug sellers

We will also select 10 drug sellers/ pharmacies per SA (see Table 1) for qualitative IDIs to investigate their knowledge, motivations, attitudes and practices around AB provision and AMR.

#### Qualitative IDIs with healthcare workers

In our recruitment hospitals /clinics, we will recruit trained clinical professionals using convenience sampling to investigate their knowledge, attitudes and AB provision/prescribing practices using qualitative IDIs (5 per SA, 45 in total).

#### Community Focus Groups

In each SA, age- and gender-specific community focus group discussions will be conducted (among non-study participants), selecting from a range of socio-economic status groups and focused on knowledge and attitudes towards UTIs, AMR and health seeking pathways. In total up to 24 FGDs per country will be conducted.

#### Patient and public involvement

In addition to the community focus groups conducted during pilot work, and the community inception workshops (detailed above), at the end of SA activities, we will use Community Dialogues (CDs) that bring together community members, health workers and veterinarians to a face-to-face engagement discussing the findings, and stimulate community participation and full engagement, and also identify how grassroots health workers might be used most effectively to improve ABR stewardship.

## Research questions and data analysis plan

The main research questions and corresponding analyses are designed within five inter-linked WS.

### WS1: The Therapy Landscape

This WS will investigate how ABs are provided and utilised in our study areas, in a number of interlinked ways. First, by analysing the geospatial data collected on AB providers, we will describe the spatial distribution and density of drug dispensing outlets in both formal and informal healthcare settings. Second, we will analyse the data from the mystery client studies geospatially. Third, by combining quantitative statistical analysis of the mystery client study with systematic thematic coding of the qualitative interviews, we will investigate the AB provision practices and knowledge among different AB providers. By developing an understanding of differences in the AB provision landscape and the knowledge, motivations and practices of AB sellers, we will seek to determine what factors regulate individual patient pathways to AB use.

### WS2: Pathogen

In this WS we will confirm urinary tract infection, identify the pathogenic organisms present, and determine the antimicrobial susceptibility. Urine samples from 1800 culture-confirmed UTI patients will be analysed to investigate the burden of disease and resistance. AST will be conducted on an agreed set of clinically relevant ABs along with special phenotypes such as extended spectrum beta-lactam resistance (ESBL) (see Appendix 1). The WHONET data capture and reporting software will be adopted in all SA hub laboratories, providing automated analysis of various multidrug resistant (MDR) phenotypes. Genomic DNA of the samples will be extracted and sequenced. The resulting whole genome sequence (WGS) libraries will be used to characterise the isolates and define pathogen population structures. We will identify the local and regional spread of AMR determinants and describe their evolutionary dynamics and local reservoirs among HATUA bacterial populations. These data will be linked to patient and household socio-demographics to pinpoint possible drivers of resistance at patient, hospital and household level. By comparing our collections with previously published genomes, high risk clones and their potential origins will be determined and their spread mapped across space and time.

### WS3: Patient

This WS will investigate social, structural and behavioural drivers of AMR by identifying the various patient pathways to treatment and how these intersect with the AMR process. We will summarise quantitative pathway data using longitudinal latent class analysis, and/or sequence analysis, and relate this statistically to AMR profiles at individual and community level. To identify how treatment pathways could become more clinically effective, we will combine quantitative and qualitative patient pathway data with in-depth interviews from drug sellers and doctors, to investigate sources of prescribed and non-prescribed ABs and practices, prevalence, and determinants of self-medication, and how these is constituted.

### WS4: Community

In this WS we will investigate social and community attitudes to treatment seeking, AB use and AMR and how community household level factors including hygiene practices, health related behaviour, livestock keeping practices, and the household microbiological landscape influence AMR and broader risk burdens. Questions in the household questionnaire addressesing AB use with animals and animal products will be statistically related to AMR burden in urine, faecal and environmental samples. We will also analyse relevant data gathered during patient IDIs and community focus groups, using systematic thematic coding in Nvivo to give us information about experiences of illness and localised rationales for treatment.

### WS5: Interdisciplinary synthesis

In this WS we will integrate and synthesise the data collected in WS 1-4 to explore how population and individual level behaviours and processes interact to contribute to the risk of AMR. Using the patient linked dataset, we will generate hypotheses about direct and indirect drivers of AMR using Bayesian network analysis. [31] We will use Bayesian networks to identify latent factors in different data types and then connect them with each other and key outcome variables in a heterogeneous network across all data. The network structure will identify direct and indirect influences on AMR, and Bayesian networks’ probabilistic inference will predict the probability of impact on AMR of change in different drivers. Multi-level regression will then be used to identify which of these direct and indirect drivers account for the most variance in outcome and provide numerical predictions of modifications.

## Discussion

### Ethical considerations

#### Informed consent

Written informed consent will be obtained from all participants prior to any data or specimen collection, with the exception of drug sellers taking part in the mystery client study, for whom the process would invalidate the approach. Participants will consent for questionnaire-filling/interviewing/focus groups (including audio recording), and for sample collection and analysis, and shipment of samples to third-party labs for WGS. All patient samples will be non-invasive by urine & stool collection only. The principal language of recruitment and administration of the informed consent document (ICD) and the questionnaire will be the language used in hospitals (i.e. Kiswahili for Kenya and Tanzania, and Luganda, Runyankole and Ngakarimojong for Uganda). Local translators will be used to draft the ICD, and ICD will be back -translated. The consent process will be administered by fieldworkers who understand the relevant languages and dialects.

#### Privacy and confidentiality

A number of procedures will be used to protect the confidentiality of respondents and the information collected: (1) Questionnaires/interviews will be conducted only in a private setting; (2) All interviewers will be trained in research ethics; (3) All data will be kept strictly confidential and numeric IDs will be used in place of names on all of the data collection instruments. At each new step of data collection, participants will be informed of confidentiality procedures during the consent process. All patients in the linked part of the study will be anonymised and identified through an 8-figure identifier, and barcode to link social science and laboratory data effectively. Any personal data will be stored on handwritten consent forms securely, and separately from questionnaire, test results and patient IDIs. EpiCollect data are uploaded to a secure cloud server. The reason for recruitment to the household follow-up will not be disclosed or discussed with anyone except the patient/respondent themselves. Geospatial data identifying home locations and drug sellers will be edited to avoid disclosure. Participants in other IDI and FGDs together with data from mystery client visits will be anonymised and recordings, translations and transcriptions will be stored and transferred securely.

#### Ethics approvals

The study received ethical approval from the University of St Andrews, UK (No. MD14548, 10/09/19); National Institute for Medical Research, Tanzania (No. 2831, updated 26/07/19), CUHAS/BMC research ethics and review committee (No. CREC /266/2018, updated on 02/2019), Mbeya Medical Research and Ethics Committee (No. SZEC-2439/R.A/V.1/303030), Kilimanjaro Christian Medical College, Tanzania (No. 2293, updated 14/08/19); Uganda National Council for Science and Technology (No. HS2406, 18/06/18); Makerere University, Uganda (No. 514, 25/04/18); and Kenya Medical Research Institute (04/06/19); Scientific and Ethics review committee(SERU) No. KEMRI/SERU/CMR/P00112/3865 V.1.2. For Uganda, administrative letters of support were obtained from the District Health Officers (DHOs) to allow the research to be conducted in the respective hospitals and health centres.

## Results dissemination, and impact

HATUA’s data will be available via an interactive website that will collate and present AMR data from across the EAC overlaid with socio-economic, microbiological and genome data. Data will be visualised and shared in Microreact (https://microreact.org) [32]. The consortium is partnered with the East African Health Research Commission (EAHRC), a statutory organ of the EAC (East African Community) which will spearhead the integration of HATUA outputs into policy at the EAC level. In the near-term, this will allow doctors to access information they can use to improve diagnosis and prescription patterns based on resistance profiles prevailing locally. In the longer–term, HATUA will lay a strong foundation for a regional surveillance initiative, and will provide a vital resource for regional AMR policy formulation. The results have great potential to inform policy, improve clinical practice and build capacity for pathogen surveillance in the region. The novel linked microbiological, genomic and social science linked data will provide new insights into social drivers of AMR.

## Data Availability

We intend to make anonymised data available for research purposes in a secure platform in due course.

## Acknowledgements

We would like to thank Graeme Sandeman for assistance with drawing the Figures.

## Authorship Statement

The following are members of the HATUA (Holistic Approach to Unravel Antibacterial Resistance in East Africa) Consortium:

Matthew T. G. Holden (UK, project PI), Benon B. Asiimwe (Co-I, Uganda), John Kiiru (Co-I, Kenya), Stephen E. Mshana (Co-I, Tanzania), Stella Neema (Co-I, Uganda), Katherine Keenan (Co-I, UK), Mike Kesby (Co-I, UK), Joseph R. Mwanga (Co-I, Tanzania), Derek J. Sloan (Co-I, UK), Blandina T. Mmbaga (Co-I, Tanzania), V Anne Smith (Co-I, UK), Stephen Gillespie (Co-I, UK), Andy G. Lynch (Co-I, UK), Alison Sandeman (UK), John Stelling (Co-I, USA), Alison Elliott (Co-I, Uganda and UK), David Aanensen (Co-I, UK, Gibson S. Kibiki (Co-I, Burundi), Wilber Sabiiti (Co-I, UK), Catherine Kansiime (Uganda), Martha F. Mushi (Tanzania), Arun Gonzales Decano (UK), Dominique L. Green (UK), John Mwaniki (Kenya), Nyanda E Ntinginya (Uganda), Joel Bazira (Uganda).

## Contributorship statement

MH led the conceptualisation of the project, helped designed protocols and data collection tools, and is the guarantor of the project. BBA contributed to the conceptualisation of the project, helped designed the tools, and led the pilot data collection and main data collection in Uganda. JK contributed to the conceptualisation of the project, helped designed the tools, and led the pilot data collection and main data collection in Kenya. SEM contributed to the conceptualisation of the project, helped designed the tools, and led the pilot data collection and main data collection in Tanzania. SN contributed to the conceptualisation of the project, helped designed the social science tools, led the pilot data collection and coordinates social science data collection in Uganda, and leads WS4. KK contributed to the conceptualisation of the project, helped designed the social science tools, contributed to the analysis plan, leads WS5, and wrote the first draft of this protocol. MK contributed to the conceptualisation of the project, helped designed the social science tools, contributed to the analysis plan. JRM contributed to the conceptualisation of the project, helped designed the social science tools, and coordinates social science data collection in Tanzania. DJS contributed to the conceptualisation of the project, helped designed the tools, and leads WS3. BTM contributed to the conceptualisation of the project, and helps coordinates data collection in Kilamanjaro, Tanzania. VAS contributed to the conceptualisation of the project, wrote parts of the data analysis plan and supervises analyses. SG contributed to the conceptualisation of the project, leads WS2, and oversees microbiological data quality. AS coordinates data collection and work stream activities and helped write this draft protocol. JS contributed to the conceptualisation of the project, and provides oversight to WS2. AE contributed to the conceptualisation of the project, and provides analytical oversight to WS2. DA contributed to the conceptualisation of the project, and the genomic analysis for WS2. GSK contributed to the conceptualisation of the project, and facilitates policy dissemination in EAC. WS contributed to the conceptualisation of the project, helped design the tools, and leads WS1. All authors revised the draft paper.

---

**Appendix A.**
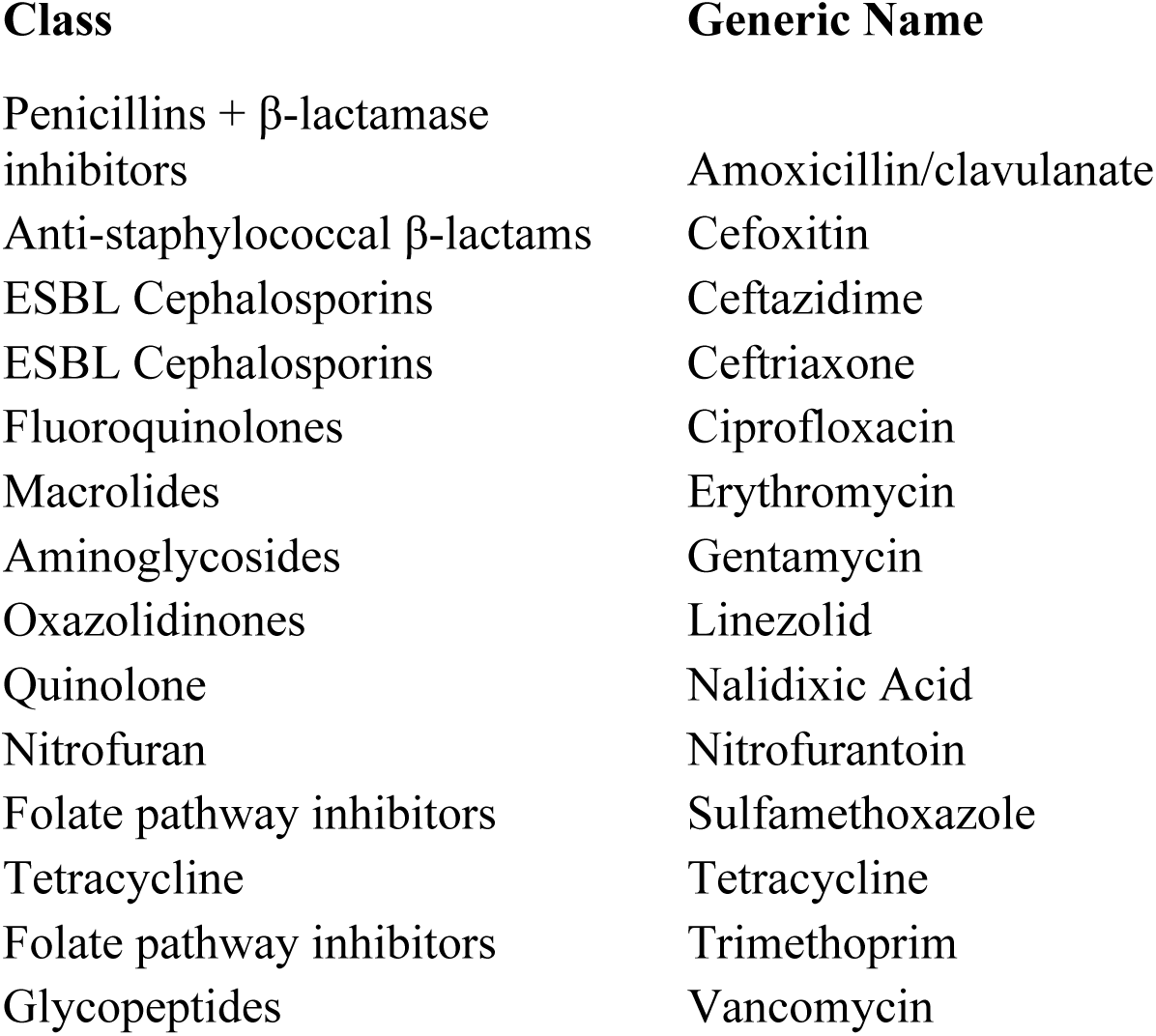
Agreed set of clinically relevant ABs for AST.

## Footnotes

1 Antibiotics in this study is specifically used to refer to drugs with antibacterial activity.

## References

1. O’Neill J. Tackling drug-resistant infections globally: final report and recommendations. London: 2016.

2. James SL, Abate D, Abate KH, et al. Global, regional, and national incidence, prevalence, and years lived with disability for 354 Diseases and Injuries for 195 countries and territories, 1990-2017: A systematic analysis for the Global Burden of Disease Study 2017. Lancet 2018;392:1789–858. doi:10.1016/S0140-6736(18)32279-7

3. Vedadhir AA, Rodrigues C, Lambert H. Social science research contributions to antimicrobial resistance: Protocol for a scoping review. Syst Rev 2020;9:24. doi:10.1186/s13643-020-1279-y

4. Haenssgen MJ, Charoenboon N, Zaw YK. It is time to give social research a voice to tackle antimicrobial resistance? J Antimicrob Chemother 2018;73:1112–3. doi:10.1093/jac/dkx533

5. Andersson DI, Hughes D. Persistence of antibiotic resistance in bacterial populations. FEMS Microbiol Rev 2011;35:901–11. doi:10.1111/j.1574-6976.2011.00289.x

6. Smith R. Antimicrobial resistance is a social problem requiring a social solution. BMJ 2015;350:h2682. doi:10.1136/bmj.h2682

7. Flores-Mireles AL, Walker JN, Caparon M, et al. Urinary tract infections: Epidemiology, mechanisms of infection and treatment options. Nat. Rev. Microbiol. 2015;13:269–84. doi:10.1038/nrmicro3432

8. Masika WG, O’Meara WP, Holland TL, et al. Contribution of urinary tract infection to the burden of febrile illnesses in young children in rural Kenya. PLoS One 2017;12:e0174199. doi:10.1371/journal.pone.0174199

9. Davis A, Sharp J. Rethinking One Health: emergent human, animal and environmental assemblages. Soc Sci Med 2020;:113093. doi:10.1016/j.socscimed.2020.113093

10. Carey G, Malbon E, Carey N, et al. Systems science and systems thinking for public health: A systematic review of the field. BMJ Open. 2015;5:e009002. doi:10.1136/bmjopen-2015-009002

11. Deleuze G, Guattari F. A Thousand Plateaus: Capitalism and Schizophrenia. 1988.https://books.google.co.uk/books?hl=en&lr=&id=5D6tAwAAQBAJ&oi=fnd&pg=PR5&dq=Deleuze,+G.,+%26+Guattari,+F.+(1988).+A+thousand+plateaus:+Capitalism+and+schizophrenia.+Bloomsbury+Publishing.&ots=ykFKkSW3Lc&sig=cAgWVgl2tLeg54i8xxTW4qs-_Lo#v=onepage&q=Deleuze%25 (accessed 10 Apr 2020).

12. Bennett J. Vibrant Matter a political ecology of things. Duke University Press. 2010. doi:10.3917/rac.025.0839

13. Susan Yi Sencindiver. New Materialism. In: Encyclopedia of Educational Philosophy and Theory. 2017. 1565–1565. doi:10.1007/978-981-287-588-4_100690

14. Bürgmann H, Frigon D, Gaze WH, et al. Water and sanitation: An essential battlefront in the war on antimicrobial resistance. FEMS Microbiol Ecol 2018;94. doi:10.1093/femsec/fiy101

15. Cox JA, Vlieghe E, Mendelson M, et al. Antibiotic stewardship in low- and middleincome countries: the same but different? Clin. Microbiol. Infect. 2017;23:812–8. doi:10.1016/j.cmi.2017.07.010

16. Morgan DJ, Okeke IN, Laxminarayan R, et al. Non-prescription antimicrobial use worldwide: a systematic review. Lancet Infect Dis 2011;11. doi:10.1016/S1473-3099(11)70054-8

17. Sakeena MHF, Bennett AA, McLachlan AJ. Non-prescription sales of antimicrobial agents at community pharmacies in developing countries: a systematic review. Int. J. Antimicrob. Agents. 2018;52:771–82. doi:10.1016/j.ijantimicag.2018.09.022

18. Ocan M, Obuku EA, Bwanga F, et al. Household antimicrobial self-medication: A systematic review and meta-analysis of the burden, risk factors and outcomes in developing countries. BMC Public Health. 2015;15:1–11. doi:10.1186/s12889-015-2109-3

19. Pescosolido BA. Beyond Rational Choice: The Social Dynamics of How People Seek Help. Am J Sociol 1992;97:1096–138. doi:10.1086/229863

20. Leventhal H, Brissette I, Leventhal EA. The common-sense model of self-regulation of health and illness. In: The Self-Regulation of Health and Illness Behaviour. Routledge 2012. 42–65. doi:10.4324/9780203553220-10

21. Chandler C, Hutchinson E, Hutchison C. Addressing Antimicrobial Resistance Through Social Theory: An Anthropologically Oriented Report. Technical Report. London: 2016.

22. Marie Stopes International, IPAS. A woman’s pathway through a medical abortion from a pharmacy. https://www.safeaccesshub.org/access/a-womans-pathway-through-a-medical-abortion-from-a-pharmacy/ (accessed 11 Apr 2020).

23. Coast E, Norris AH, Moore AM, et al. Trajectories of women’s abortion-related care: A conceptual framework. Soc. Sci. Med. 2018;200:199–210. doi:10.1016/j.socscimed.2018.01.035

24. Coast E, Murray SF. ‘These things are dangerous’: Understanding induced abortion trajectories in urban Zambia. Soc Sci Med 2016;153:201–9. doi:10.1016/j.socscimed.2016.02.025

25. Haenssgen MJ, Ariana P. Healthcare access: A sequence-sensitive approach. SSM - Popul Heal 2017;3:37–47. doi:10.1016/J.SSMPH.2016.11.008

26. Haenssgen MJ, Charoenboon N, Zanello G, et al. Antibiotics and activity spaces: protocol of an exploratory study of behaviour, marginalisation and knowledge diffusion. BMJ Glob Heal 2018;3:e000621. doi:10.1136/bmjgh-2017-000621

27. Risso-Gill I, Balabanova D, Majid F, et al. Understanding the modifiable health systems barriers to hypertension management in Malaysia: A multi-method health systems appraisal approach. BMC Health Serv Res 2015;15. doi:10.1186/s12913-015-0916-y

28. Decano AG, Pettigrew K, Sabiiti W, et al. Pan-resistome characterization of uropathogenic Escherichia coli and Klebsiella pneumoniae strains circulating in Uganda and Kenya isolated from 2017-2018. medRxiv Prepr Published Online First: 2020. doi:10.1101/2020.03.11.20034389

29. Aanensen DM, Huntley DM, Feil EJ, et al. EpiCollect: Linking smartphones to web applications for epidemiology, ecology and community data collection. PLoS One 2009;4. doi:10.1371/journal.pone.0006968

30. Wafula FN, Miriti EM, Goodman CA. Examining characteristics, knowledge and regulatory practices of specialized drug shops in Sub-Saharan Africa: A systematic review of the literature. BMC Health Serv. Res. 2012;12:223. doi:10.1186/1472-6963-12-223

31. Heckerman D, Geiger D, Chickering DM. Learning Bayesian networks: The combination of knowledge and statistical data. Mach Learn 1995;20:197–243. doi:10.1007/BF00994016

32. Argimón S, Abudahab K, Goater RJE, et al. Microreact: visualizing and sharing data for genomic epidemiology and phylogeography. Microb genomics 2016;2:e000093. doi:10.1099/mgen.0.000093

